# Electroconvulsive therapy increases cortical thickness in depression: A systematic review

**DOI:** 10.1101/2023.10.22.23297375

**Authors:** Toffanin Tommaso, Cattarinussi Giulia, Ghiotto Niccolò, Lussignoli Marialaura, Pavan Chiara, Pieri Luca, Schiff S., Finatti Francesco, Romagnolo Francesca, Folesani Federica, Nanni Maria Giulia, Caruso Rosangela, Zerbinati Luigi, Belvederi Murri Martino, Ferrara Maria, Pigato Giorgio, Grassi Luigi, Sambataro Fabio

## Abstract

**Objective:** Electroconvulsive therapy (ECT) is one of the most studied and validated available treatments for severe or treatment-resistant depression. However, little is known about the neural mechanisms underlying the ECT treatment. This systematic review aims to critically review all structural magnetic resonance imaging studies investigating longitudinal cortical thickness (CT) changes after ECT in patients with unipolar or bipolar depression.

**Methods:** We performed a search on PubMed, Medline, and Embase to identify all available studies published before April 20, 2023. A total of 10 studies were included.

**Results:** The investigations showed widespread increases in CT after ECT in depressed patients, involving mainly the temporal, insular, and frontal regions. In five studies, CT increases in a non-overlapping set of brain areas correlated with the clinical efficacy of ECT. The small sample size, heterogeneity in terms of populations, medications, comorbidities, and ECT protocols, and the lack of a control group in some investigations limit the generalizability of the results.

**Conclusions:** Our findings support the idea that ECT can increase CT in patients with unipolar and bipolar depression. It remains unclear whether these changes are related to the clinical response. Future larger studies with longer follow-up are warranted to thoroughly address the potential role of CT as a biomarker of clinical response after ECT.

**Summations:**

- This review summarizes how ECT affects CT in patients with unipolar or bipolar depression.
- The areas that were predominantly affected by ECT were temporo-insular and frontal regions. An association between the antidepressant effect of ECT and CT changes was reported by half of the included studies.
- Identifying the possible cortical changes associated with the clinical efficacy of ECT opens new targets to ameliorate ECT protocols.

**Considerations:** The review is based on studies with small numbers of patients and considerable heterogeneity in terms of patients’ characteristics and ECT protocols. Most studies cited did not have a randomized design, thus reducing the strength of evidence supporting a causal link between ECT and CT changes.

## Introduction

Electroconvulsive therapy (ECT) was first used in 1938 by Italian scientists Ugo Cerletti and Lucio Bini to induce an epileptic seizure to treat patients with schizophrenia (Gazdag and Ungvari, 2019). In the following decades, the introduction of general anesthesia and accurate operational protocols greatly improved ECT techniques (Kadiyala and Kadiyala, 2018; Kellner et al., 2020), allowing ECT to evolve to become a promising and well-tolerated treatment. Nowadays, it is considered the safest procedure performed under general anesthesia (Tørring et al., 2017). ECT is an important therapeutic strategy for affective disorders, including severe major depressive disorder (MDD),particularly with psychotic symptoms and a high risk of suicide, treatment-resistant depression (TRD), and depression in bipolar disorder (BD) (Fornaro et al., 2020; Kho et al., 2003). Despite the major advances in psychopharmacology and brain stimulation techniques, ECT remains one of the most effective treatments in psychiatry, with response rates in the treatment of unipolar and bipolar depression ranging from 74 to 77 % (Bahji et al., 2019).

The exact pathophysiological mechanisms underlying depression have not yet been clarified. Among the putative mechanisms, the “neurotrophin hypothesis of depression” suggests that a stress-induced decrease in the expression of brain-derived neurotrophic factor (BDNF) leads to atrophy of stress-vulnerable neurons in the hippocampus (Duman et al., 1997). Later studies expanded this theory, indicating that changes in BDNF expression and function result in structural brain alterations not only in the hippocampus but also in the prefrontal cortex (PFC) (Duman and Li, 2012). Crucially, magnetic resonance imaging (MRI) investigations have been supporting this theory, demonstrating reductions in gray matter volumes in MDD in the hippocampus and orbitofrontal cortex, as well as in the basal ganglia, thalamus, and anterior cingulate cortex (ACC)(Kempton et al., 2011; Koolschijn et al., 2009). In bipolar depression, similar volumetric alterations have been reported, particularly in the frontal and limbic areas. In addition to volumetric changes, alterations in surface measurements have also been reported in depression, such as cortical thickness (CT), which represents the depth of gray matter as the covering neural sheet of brain foldings and is correlated with the size of neurons, neuroglia, and nerve fibers (Narr et al., 2005). Importantly, a reduction in CT could indicate a reduction in dendritic arborization or changes in myelination at the gray/white matter interface Fare clic o toccare qui per immettere il testo.(Sowell et al., 2004) The Enhancing Neuroimaging Genetics through Meta-Analysis (ENIGMA) Consortium demonstrated that lower CT in temporal regions is a common feature in various neuropsychiatric disorders (Boedhoe et al., 2018; Hoogman et al., 2019; Schmaal et al., 2017; Van Rooij et al., 2018). With regards to MDD, the ENIGMA Major Depressive Disorder Working Group reported thinner cortical gray matter in MDD compared to controls in the orbitofrontal cortex (OFC), ACC and posterior cingulate cortex (PCC), insula, and temporal lobes (Schmaal et al., 2017). Furthermore, a decrease in CT was also found in elderly patients with MDD (Lim et al., 2012) and in those with a high risk for suicide (Wagner et al., 2012) Additionally, a thinner CT in the PCC could indicate a higher risk of nonremission in MDD patients (Järnum et al., 2011). In BD, a recent ENIGMA study highlighted that individuals with BD showed a widespread thinner cortex relative to HC, with the largest effect size in the left fusiform gyrus (Matsumoto et al., 2023).

Although the association between surface measures and the severity of depressive symptoms is unclear, changes in CT after response and remission have been reported after antidepressant treatment. In particular, in a 12-week trial using sertraline in patients with late-life depression, nonremitters presented thinner frontal poles at baseline compared to remitters (Sheline et al., 2012). Another trial showed that sertraline increased CT in the left medial PFC, right medial and lateral PFC, and within the right parietal-temporal lobes in patients with MDD compared to placebo (Nemati and Abdallah, 2020). In addition, CT also predicted the effectiveness of psychotherapy for late-life depression (Mackin et al., 2013). Therefore, reduced CT appears to be related to the reduced efficacy of pharmacological and pharmacotherapy interventions in MDD, and successful treatment appears to reverse atrophy in fronto-limbic areas (Motter et al., 2021). Crucially, reduced CT appears to be associated also with early-life traumas and with other affective disorders, including BD and anxiety (Yang et al., 2023; Zhu et al., 2022).

The neural mechanisms underlying the effects of ECT in the treatment of depression are still largely unknown, and this further contributes to the skepticism about its use (Bolwig, 2011). Importantly, recent meta-analytic evidence suggested that ECT is associated with volume increases in the hippocampus and increased integrity of white matter pathways in the frontal and temporal lobes(Gbyl and Videbech, 2018) These findings have also been confirmed by a mega-analysis conducted by the Global ECT-MRI Research Collaboration (GEMRIC), which showed that ECT induced broadly distributed gray matter volumetric increases (Ousdal et al., 2020). Interestingly, CT may be more sensitive than volume to brain changes associated with ECT, including alterations in the glial and neuronal microstructure (Winkler et al., 2010).

Given these premises, we aimed to review the available evidence on CT alterations associated with ECT treatment in patients with depression to explore the association between changes in CT and the antidepressant effect of ECT.

## Materials and methods

### Literature search and screening

The study inclusion criteria were the following: a) include participants with a diagnosis of MDD or BD according to the Diagnostic and Statistical Manual of Mental Disorders (DSM) or International Classification of Disease (ICD) criteria or TRD (any definition); b) participants underwent MRI pre- and post-ECT treatments; c) changes of CT were assessed longitudinally, i.e. before and after the ECT treatments; c) CT changes were longitudinally evaluated, i.e., before and after the ECT sessions (no restrictions in terms of the number of sessions or temporal distance between ECT and MRI).

A comprehensive search of the literature was performed on PubMed, Medline, and Embase using the following keywords: (ECT [Title/Abstract] OR Electroconvulsive Therapy [Title/Abstract] OR Electroconvulsive Treatment [Title/Abstract]) AND ((Major Depressive Disorder [Title/Abstract] OR Depression [Title/Abstract] OR Bipolar Disorder [Title/Abstract]) OR Bipolar Depression [Title/Abstract]) AND Cortical thickness [Title/Abstract]). No language restrictions were applied to identify peer-reviewed articles published up to April 2023.

We screened titles and abstracts of all potentially eligible studies, retrieving the full-text articles. References cited by each study were also manually screened to ensure that no relevant studies were left out. This procedure was carried out according to the Preferred Reporting Items for Systematic Reviews and Meta-Analysis (PRISMA) guidelines (Liberati et al., 2009). Of the 35 initial studies, 25 remained after removing duplicates, and 11 citations met the title/abstract screening eligibility criteria and were included. After full-text screening, 1 article was excluded because it did not report longitudinal changes after ECT (Wade et al., 2017), resulting in 10 eligible studies (Fig.1). Since only two studies reported quantitative measures of the effects of ECT on CT, we could not perform a meta-analysis.

**Fig. 1.**
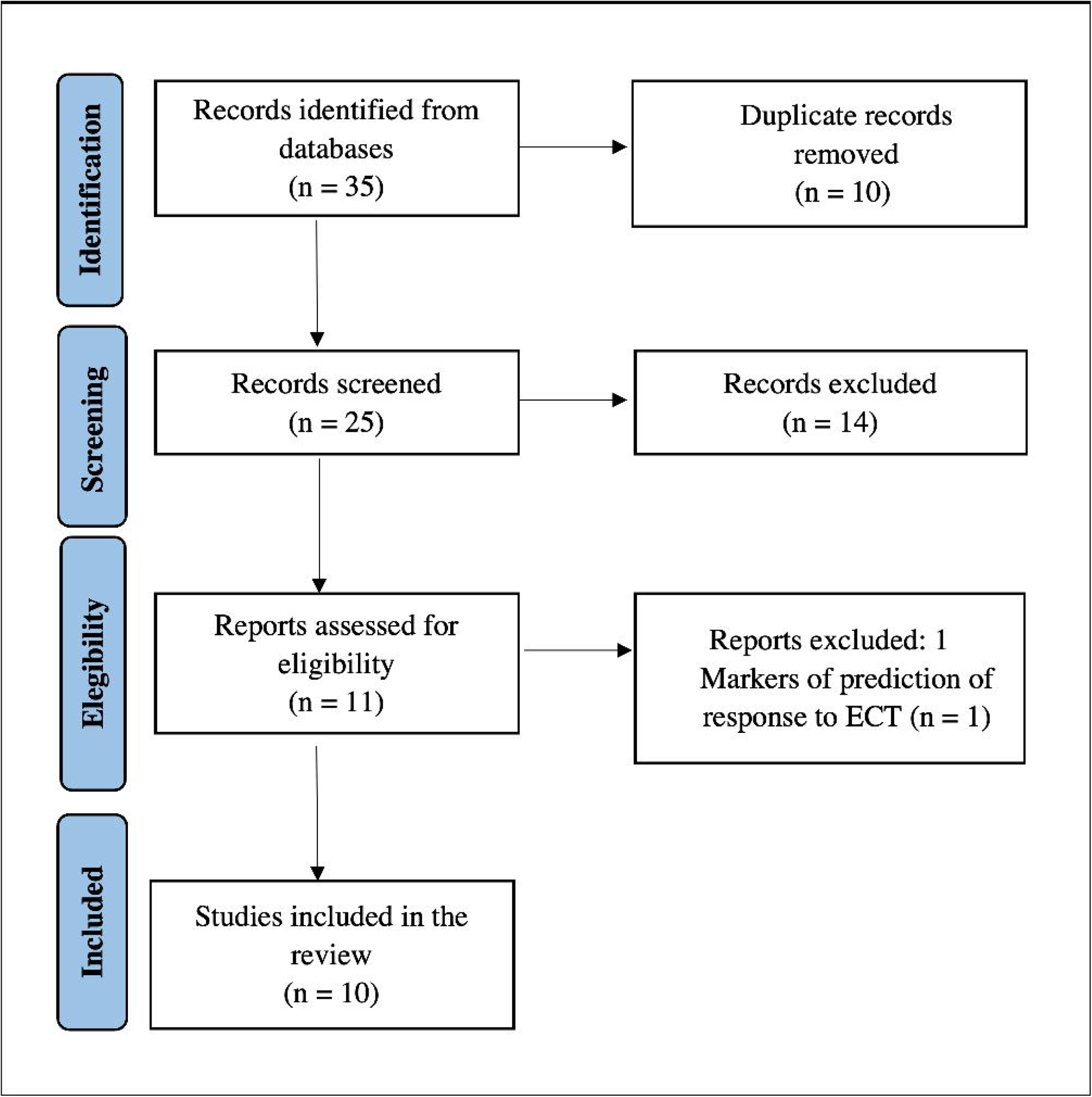
PRISMA flow chart of selection of publications for inclusion in the review.

### Assessment of methodological quality

The methodological quality of the included studies was assessed using a point system derived from Gbly and Videbech (Gbyl and Videbech, 2018). Briefly, the quality score consisted of the following items: a) number of subjects; b) presence of a control group; c) number of MRI scans the control group underwent; d) MRI scanner field strength; e) voxel size of the MRI scan; f) medication status; g) consecutively collected sample; h) duration of follow-up. A higher score indicated better methodological quality (see Supplementary Material).

## Results

### Demographic, clinical, and ECT characteristics of included studies

The characteristics of the included studies are described in Tab. 1. 253 patients (244 with MDD or TRD, 9 with bipolar depression, mean age 46.3 ± 10.6 years; 67.9 % female; mean duration of the illness 17.7 years) and 143 healthy controls (mean age 48.9 ± 11.2 years; 51.7 % female) were included. Seven studies reported that depressed participants received medications at the time of scan (Bracht et al., 2023; Gbyl et al., 2019; Gryglewski et al., 2019; Sartorius et al., 2016; Schmitgen et al., 2020; Xu et al., 2019; Yrondi et al., 2019). Seven studies repeated structural analysis twice (Bracht et al., 2023; Gryglewski et al., 2019; Ji et al., 2022; Sartorius et al., 2016; Schmitgen et al., 2020; van Eijndhoven et al., 2016; Xu et al., 2019) and three studies performed three MRI scans (Gbyl et al., 2019; Pirnia et al., 2016; Yrondi et al., 2019). In all studies, the first scan was acquired within a week before the ECT session and the second within a week after treatment. Two studies acquired an intermediate scan between the first and last ECT sessions to explore acute changes in CT (Pirnia et al., 2016; Yrondi et al., 2019), while one study acquired a scan before the first ECT session, a scan after the last ECT treatment, and a scan within six months after ECT treatment to examine the long-lasting effects of ECT on CT (Gbyl et al., 2019) (Fig. 2). The diagnosis of MDD was made according to DSM-IV, DSM-IV-TR, and DSM-5. Depressive symptoms were assessed with the Hamilton depression rating scales (HAM-D) (Hamilton, 1960) and in Pirnia et al. (2016) (Pirnia et al., 2016) also with the Montgomery-ALsberg depression rating scales (MADRS) (Montgomery and Asberg, 1979). In most studies, the medications were kept unchanged before ECT (Bracht et al., 2023; Gbyl et al., 2019; Gryglewski et al., 2019; Schmitgen et al., 2020; Wade et al., 2017; Xu et al., 2019), while in two studies the medications were tapered (Sartorius et al., 2016; van Eijndhoven et al., 2016). Patients were excluded when they presented neurological disorders, addictive disorders, schizophrenia spectrum disorders and personality disorders, a history of head injury resulting in unconsciousness for more than 5 minutes, MRI contraindications, and previous ECT sessions received in the last six months/year.

**Fig. 2.**
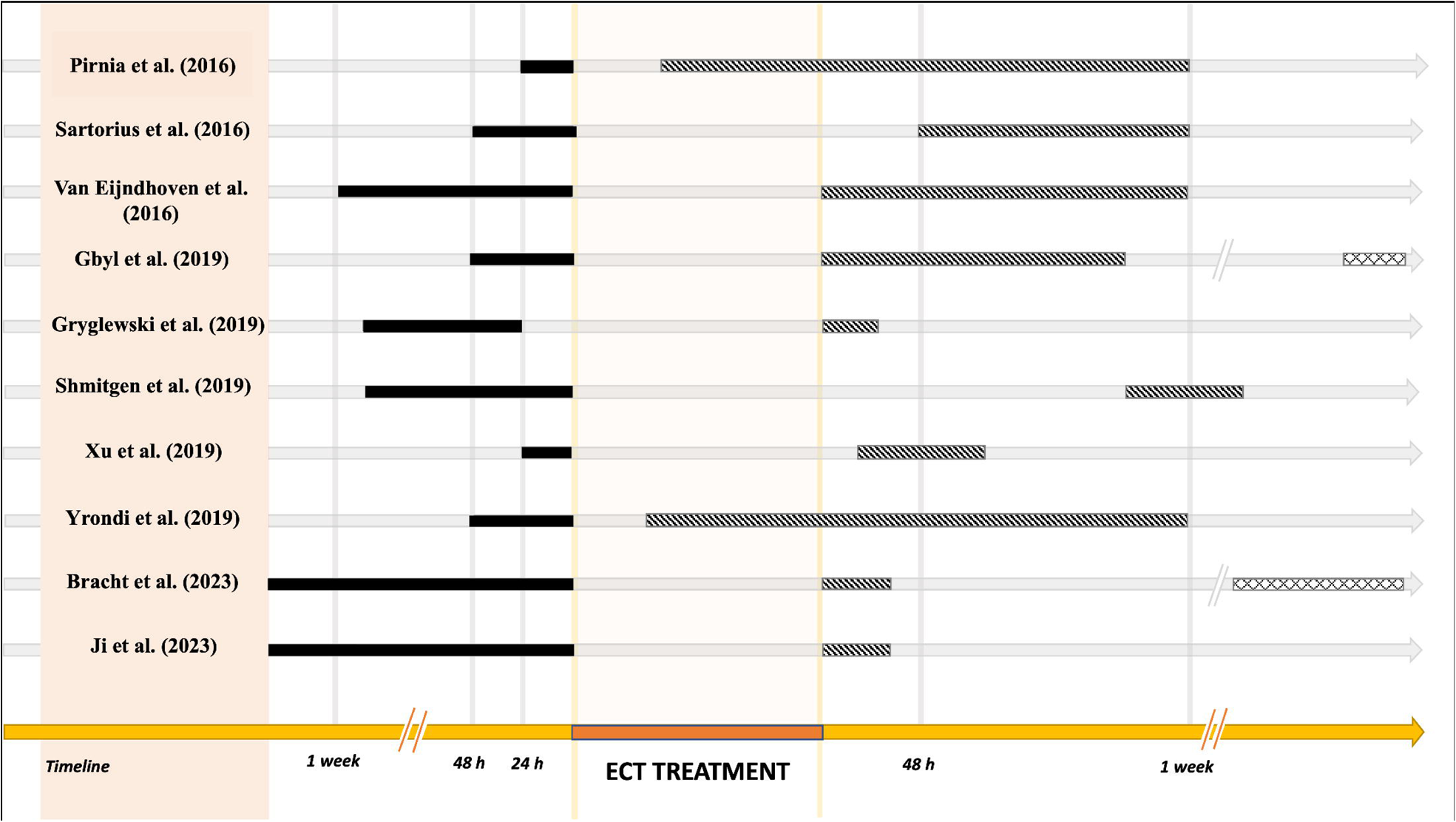
Magnetic resonance imaging (MRI) scan timeline of the studies of the effect of ECT on cortical thickness in patients with depression. For each study, we have reported the timing of the structural magnetic resonance images relative to the entire ECT treatment (in orange), including the baseline scan (solid black line, T1), the second scan (hatched line, T2), and the third scan (cross-hatched line, T3). The time is measured relative to the beginning (T1) and end (T2, T3) of the ECT treatment.

**Table 1:**
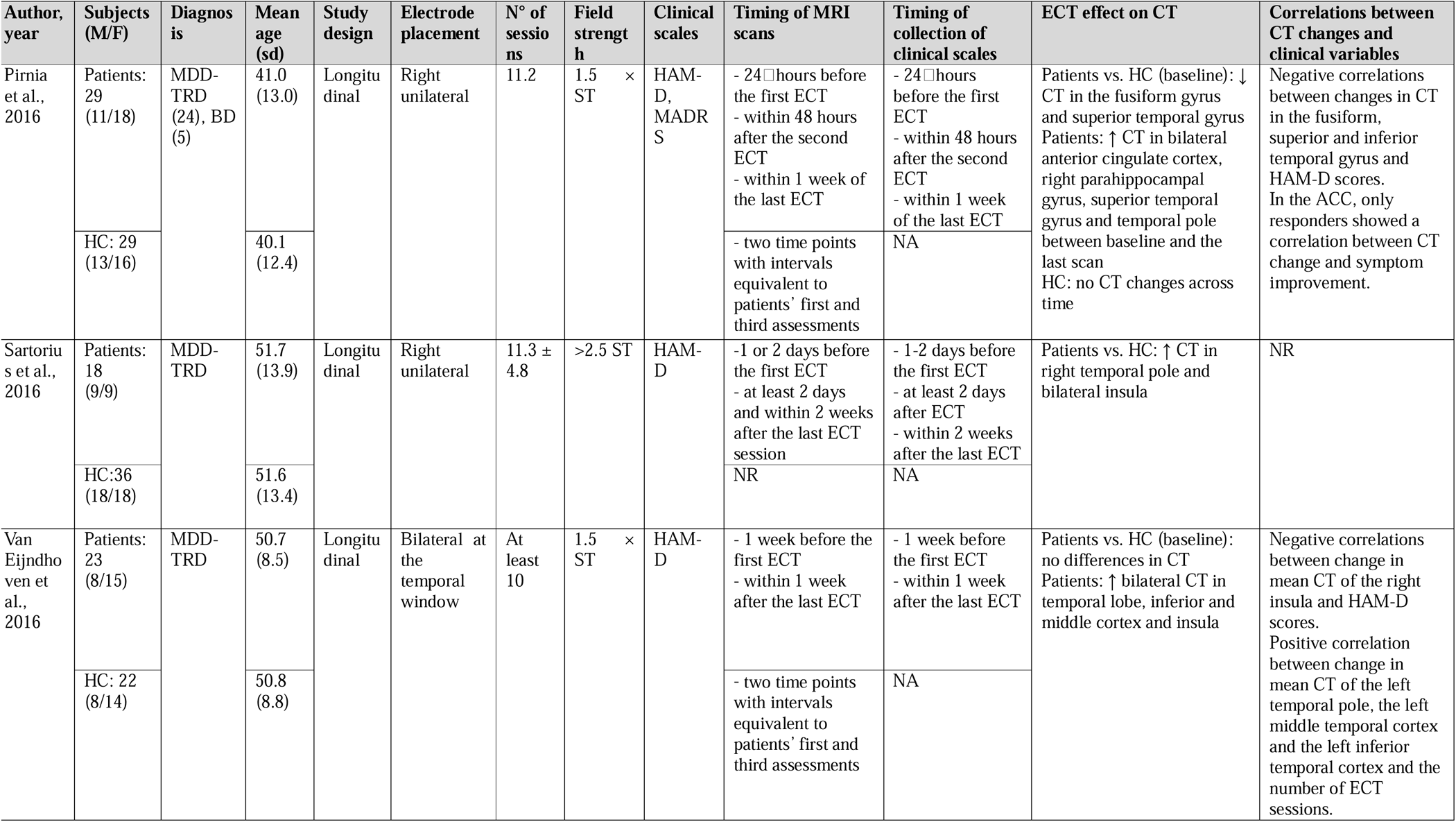
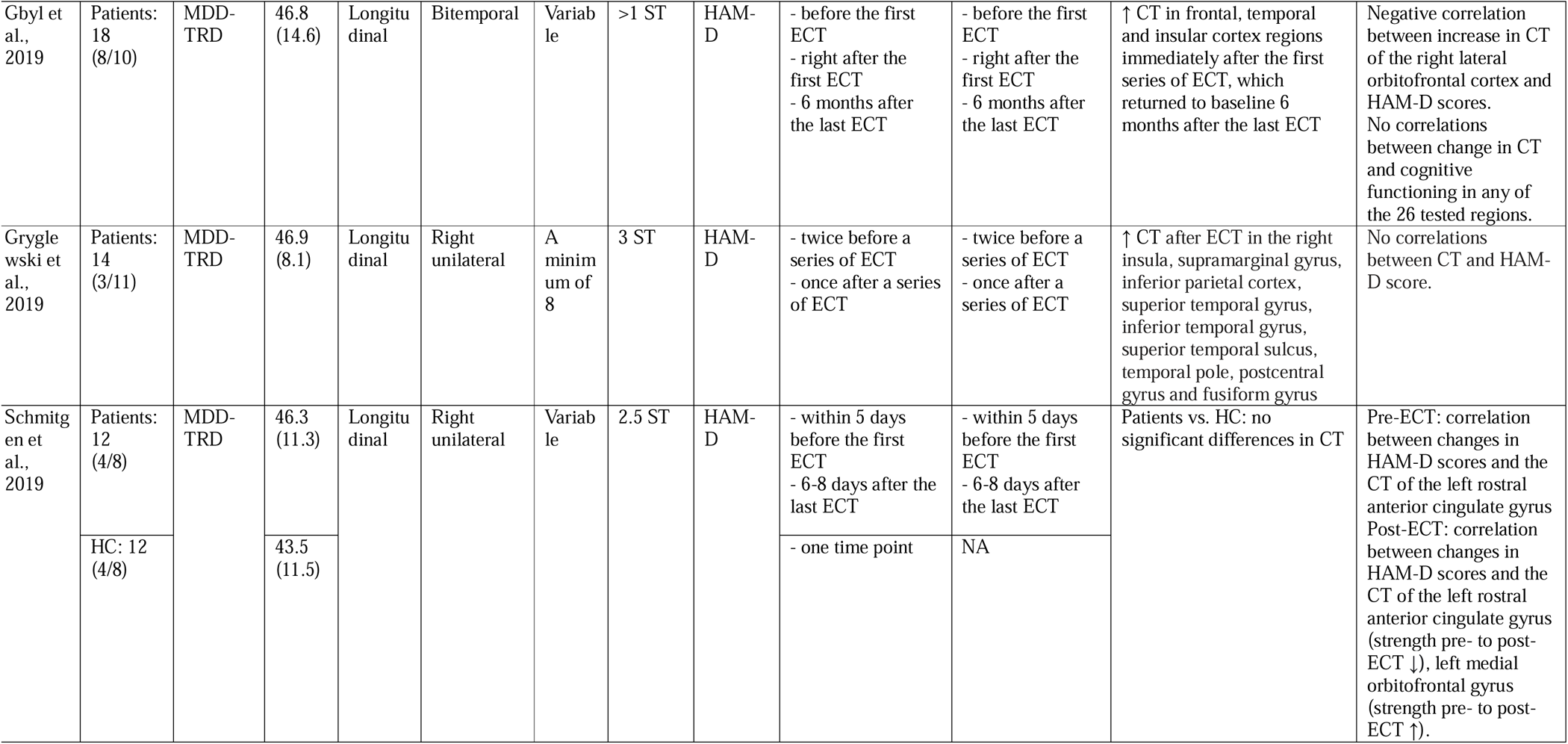
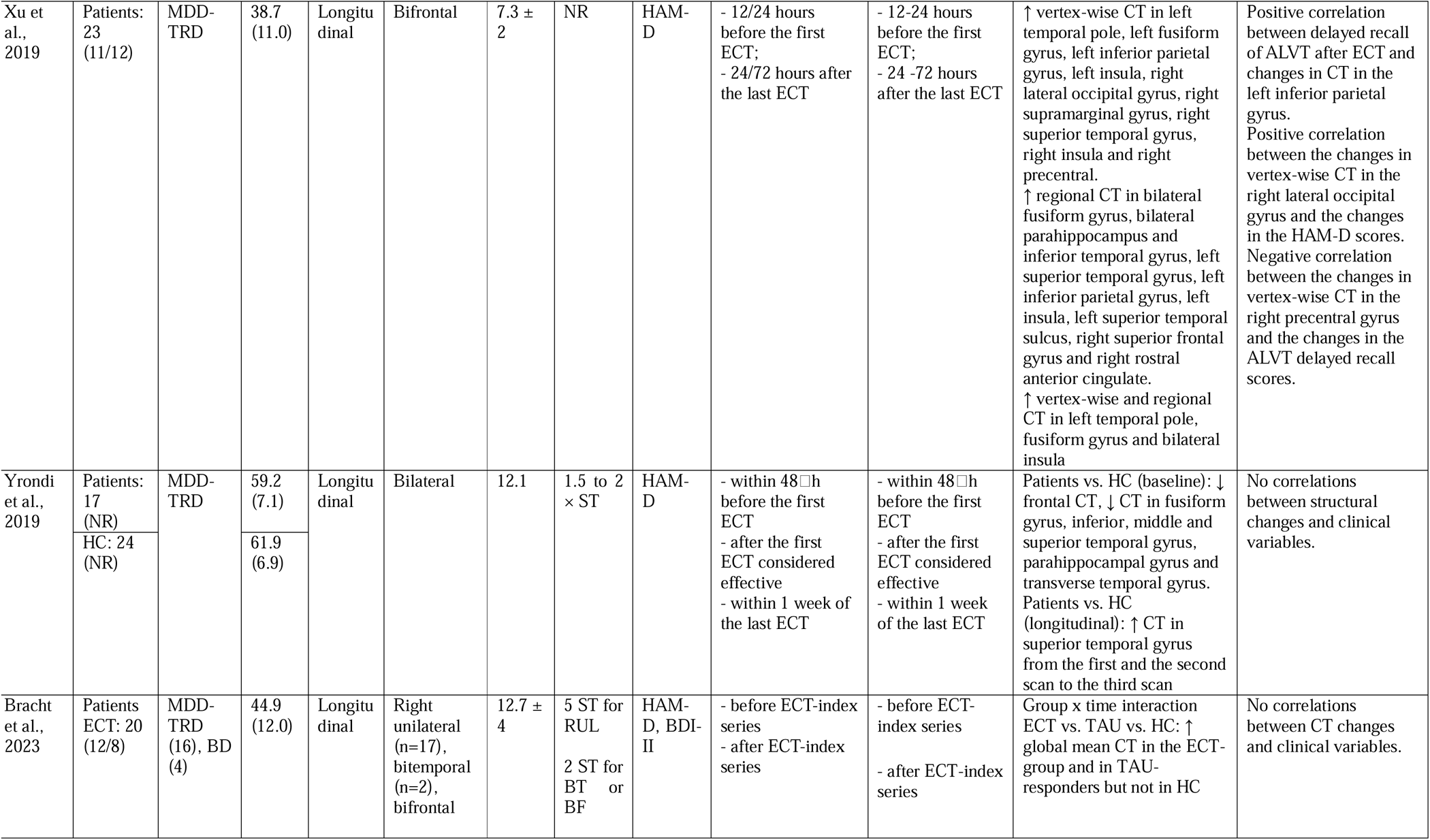
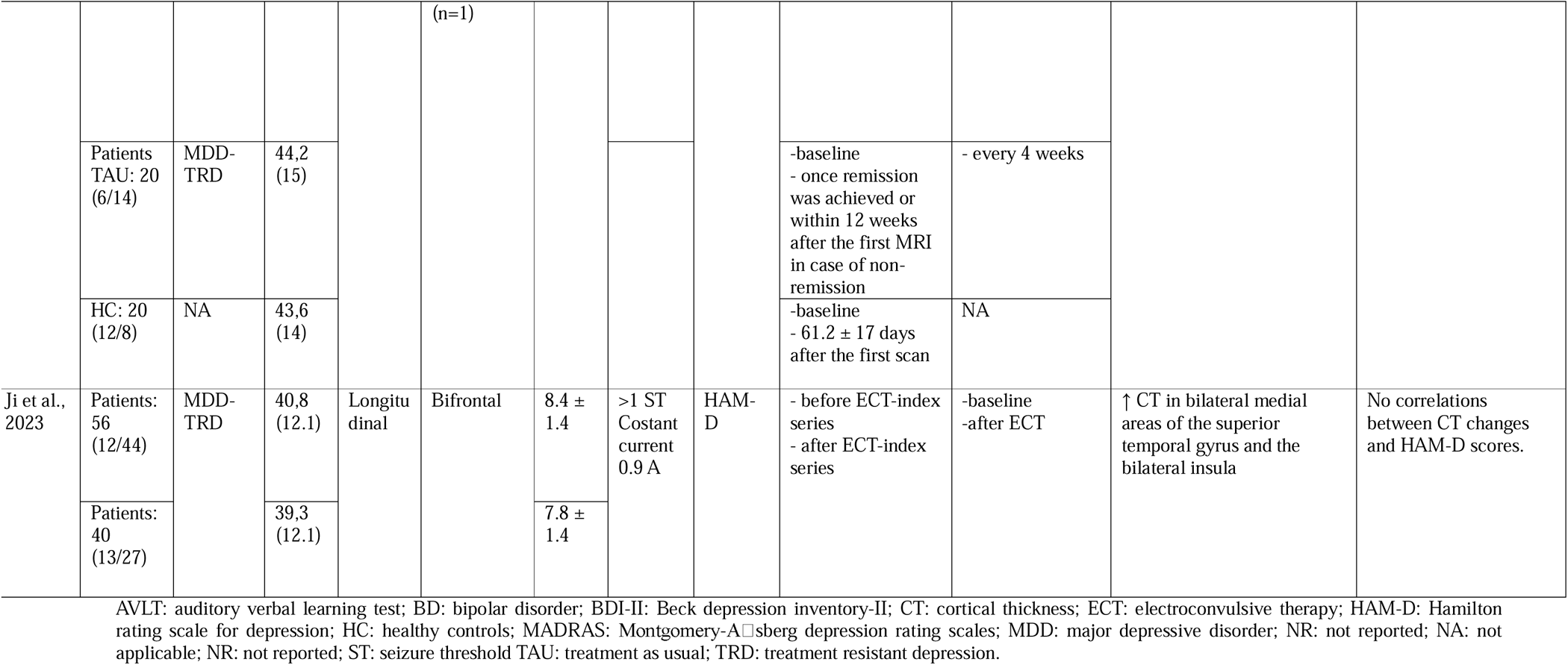
Studies investigating the effect of ECT on CT in depressed patients.

Six studies included a comparison with a group of healthy participants who did not undergo ECT treatment, while Bracht et al. (2023) also included an MDD group who received Treatment As Usual (TAU) (Bracht et al., 2023). Regarding the ECT protocols, five studies used bilateral electrode placement (Gbyl et al., 2019; Ji et al., 2022; van Eijndhoven et al., 2016; Xu et al., 2019; Yrondi et al., 2019), four studies used unilateral electrode placement (Gbyl et al., 2019; Gryglewski et al., 2019; Pirnia et al., 2016; Sartorius et al., 2016; Schmitgen et al., 2020; Wade et al., 2017). The study by Bracht et al. (2023) used mixed protocols: 17 patients started with right unilateral ECT, two patients with bitemporal ECT, and one patient with bifrontal stimulation (Bracht et al., 2023).

### CT changes after ECT

Most studies identified CT alterations in the temporal, insular and frontal areas, mainly proximally to the electrode (Fig. 3). Gryglewski et al. (2019) and Sartorius et al. (2016) combined voxel-based morphometry and CT analysis (Gryglewski et al., 2019; Sartorius et al., 2016). The first investigation reported a widespread increase in CT of the right hemisphere, including the insula, supramarginal gyrus, inferior parietal gyrus, superior temporal gyrus, inferior temporal gyrus, superior temporal sulcus, temporal pole, postcentral gyrus and fusiform (Gryglewski et al., 2019). Similarly, Sartorius et al. (2016) found an increase in CT in the right temporal pole and the bilateral insula gyrus in MDD compared to HC (Sartorius et al., 2016). Van Eijndhoven et al. (2016) revealed large bilateral clusters of increased CT in TRD after ECT treatment extending from the temporal pole, middle and superior temporal gyrus to the insula, and inferior temporal gyrus in the left hemisphere. Post-hoc analyses demonstrated that the increase in CT of the insular cortex was greater in responders compared to non-responders to ECT. Notably, a higher number of ECT sessions was correlated with greater mean CT of the left temporal pole and the left middle and inferior temporal cortex (van Eijndhoven et al., 2016). Xu et al. (2019) calculated vertex-wise and regional CT within each of the 68 anatomically defined regions from the Desikan–Killiany Atlas in MDD (Xu et al., 2019). The authors reported a higher vertex-wise CT in the left temporal pole, left fusiform gyrus, left inferior parietal gyrus, left insula, as well as in the right lateral occipital gyrus, right supramarginal gyrus, right superior temporal gyrus, right insula and right precentral gyrus in patients with MDD after ECT. Furthermore, a greater regional CT was observed in the bilateral fusiform gyrus, bilateral parahippocampus, inferior temporal gyrus, left superior temporal gyrus, left inferior parietal gyrus, left insula, left superior temporal sulcus, right superior frontal gyrus and the right anterior rostral cingulate, as well as increased vertex-wise and regional CT in the left temporal pole, the fusiform gyrus and bilateral insula in MDD patients after ECT(Xu et al., 2019). Pirnia et al. (2016) and Yrondi et al. (2019) to better delineate the course of CT changes during the entire ECT treatment, acquired an initial scan (T1), an intermediate scan between the first and the last ECT session (T2), and a scan within one week of the last ECT (T3) (Pirnia et al., 2016; Yrondi et al., 2019). In the study of Yrondi et al (2019), the second MRI was performed immediately after the first ECT was considered effective (Yrondi et al., 2019). The researchers detected a significant increase in CT in the superior temporal gyrus in the patient group between baseline and time T3, between T2 and T3, but not between T1 and T2, suggesting that CT changes occur only after multiple ECT sessions. A post-hoc analysis revealed that, at baseline, MDD had thinner CT in the frontal lobe, in the fusiform gyrus, inferior, middle, and superior temporal gyri, parahippocampal gyrus, and the transverse temporal gyrus compared to HC. These differences were no longer significant after ECT (Yrondi et al., 2019). In Pirnia et al. (2016), the second scan was performed within 48 hours after the second ECT treatment; CT became significantly thicker in the bilateral anterior cingulate cortex, the right parahippocampal gyrus, the superior temporal gyrus, and the temporal pole between T1 and T3 in patients with unipolar and bipolar depression. An additional ROI analysis showed significant effects of ECT in the anterior cingulate cortex, parahippocampal, entorhinal, superior temporal, inferior temporal, and fusiform cortex. To determine whether ECT was associated with a normalization toward control values, differences in CT between patients and HC were examined at baseline across the whole cortex and in ROIs that showed significant effects of ECT. Within the cortical ROIs, at baseline, patients showed reduced CT in the fusiform and superior temporal cortex compared to HC, which was not present at the two subsequent time points(Pirnia et al., 2016). Gbyl et al. (2019) explored CT changes in MDD from baseline to six months after the last ECT, with an intermediate scan after the first ECT. They reported an increase in CT in 26 cortical regions, mainly within the frontal, temporal, and insular cortex immediately after the first series of ECT, which returned to baseline at six months of follow-up (Gbyl et al., 2019).A recent study by Bracht et al. (2023) compared longitudinal changes in CT between the ECT-treated group, treatment-as-usual (TAU) responders, and HC. Patients with unipolar and bipolar TRD were scanned twice, at the beginning and at the end of the ECT series, while patients with TAU were scanned at baseline and after achieving remission of depressive symptoms or within 12 weeks after the first MRI scan (Bracht et al., 2023). Global mean CT was increased in the right hemisphere in the ECT group and bilaterally in the TAU responders. Exploratory analyses revealed that CT changes common to the ECT group and TAU-responders included the right insula and the right lateral orbitofrontal cortex (Bracht et al., 2023). Another recent longitudinal study explored CT in 96 patients with MDD before and after ECT and reported CT increases in bilateral medial areas of the superior temporal gyrus and bilateral insula compared to baseline (Ji et al., 2022). Lastly, Schmitgen et al. (2020) investigated several surface-based measures in 12 patients with TRD and found no differences in CT relative to 12 HC and no longitudinal changes of this parameter in the group of patients before and after ECT (Schmitgen et al., 2020).

**Fig. 3.**
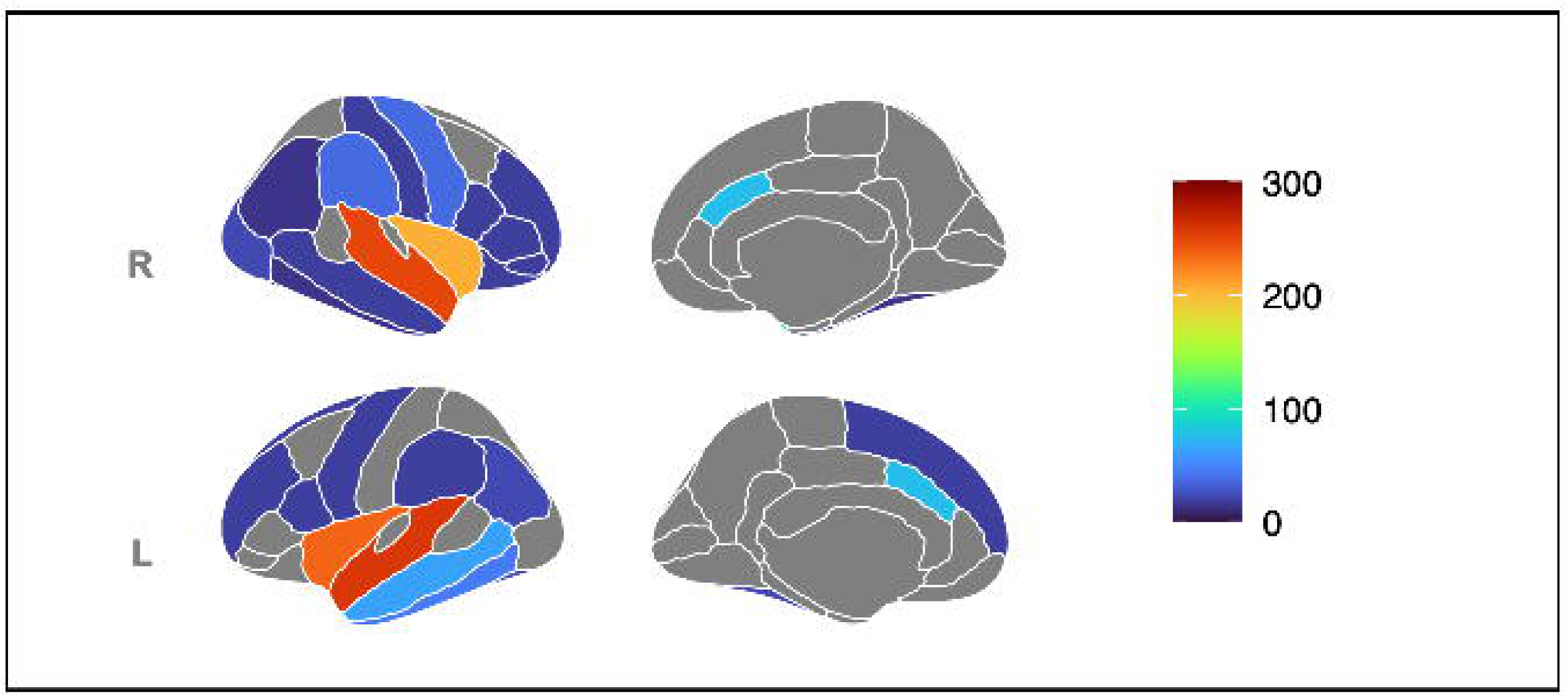
Changes in cortical thickness in patients with depression after ECT. Results from morphometric studies are displayed on the Desikan-Killiany atlas. The medial and lateral cortical surfaces are displayed for each hemisphere on the leftmost part, the subcortical regions in the center, and the colorbar on the rightmost part of each panel, respectively. The color bar code indicates the total number of subjects who showed an increase in cortical thickness. L, left hemisphere; R, right hemisphere. The renderings were created using the R-package *ggseg*.

### Correlations between CT changes and clinical improvements

Ten studies explored the correlations between CT changes and clinical improvements in depression measured with HAM-D between baseline and follow-up, and five reported a significant association. Gbyl et al. (2019) found a positive correlation between CT increases in the right lateral orbitofrontal gyrus and clinical improvement of depression (Gbyl et al., 2019). Similarly, positive correlations were observed between changes in CT in the fusiform, superior and inferior temporal gyrus and clinical improvement in depression(Pirnia et al., 2016); in particular, CT of the anterior cingulate cortex increased shortly (T1 and T2) after the initiation of ECT treatment and was predictive of the general clinical response (>50% improvement in HAM-D scores) to ECT. In van Eijndhoven et al. (2016), increased mean CT of the right insula was associated with clinical improvement in depressive symptoms (van Eijndhoven et al., 2016). Schmitgen et al. (2020) observed that pre- and post-ECT CT of the left rostral anterior cingulate gyrus predicted a greater improvement in depression. Furthermore, they also found a correlation between clinical improvement and increased CT of the left medial orbitofrontal gyrus after but not before ECT treatment, suggesting a mediator role of cortical changes in this region (Schmitgen et al., 2020). Lastly, Xu et al. (2019) reported a correlation between vertex-wise CT increases in the left occipital gyrus and the improvement of depression. Furthermore, to test the brain mechanisms of tolerability of ECT, the authors explored the association between cognitive functions and CT changes. In particular, ECT-dependent memory impairments measured by Auditory Verbal Learning Test (ALVT) delayed recall scores were negatively correlated with CT increase in the left inferior parietal gyrus and marginally in the right precentral gyrus after ECT treatment (Xu et al., 2019).

## Discussion

This review summarized the evidence on CT changes associated with ECT among patients with depression. We found that ECT was associated with a widespread increase in CT in patients with depression, located primarily in the temporal lobe and insula cortex and, to a lesser extent, in the frontal areas. Some studies showed a correlation between increases in CT and clinical improvement.

### Temporal lobe

Most of the reviewed studies found that ECT was followed by increased CT in the temporal lobe, including the superior, middle, and inferior temporal gyri and temporal pole (Gbyl et al., 2019; Gryglewski et al., 2019; Ji et al., 2022; Pirnia et al., 2016; Sartorius et al., 2016; van Eijndhoven et al., 2016; Xu et al., 2019; Yrondi et al., 2019). Although most increases were observed near the placement of the electrodes, temporal changes were also observed when the electrodes were located in the frontal lobe (Ji et al., 2022; Xu et al., 2019). Importantly, ECT has two main therapeutic components: seizure activity and electric perturbation due to an induced electric field (Fridgeirsson et al., 2021). The initial idea that the main benefits of ECT came from induced seizures has been disputed, given the evidence that increasing the strength of the electric field can affect the clinical effectiveness of ECT, without affecting the characteristics of the seizure(Argyelan et al., 2019). Notably, current literature suggests that greater intensity of the electric field may be associated with greater neuronal plasticity after treatment (Argyelan et al., 2019). In this regard, animal studies have shown that electroconvulsive seizures stimulate dendritic arborization, synaptic density, and glial proliferation (Maynard et al., 2018; Ongür et al., 2007). This may be a consequence of the increased expression and release of brain-derived neurotrophic factor induced by ECT (Polyakova et al., 2015), which could ultimately be the basis for the therapeutic efficacy of ECT. However, whether the efficacy of ECT is related to the electric field or the resulting seizure, or both, remains a debated matter (Deng et al., 2023). The temporal lobe plays a crucial role in the pathophysiology of depression, as it is involved in the core neuropsychological features that are commonly disrupted in MDD and bipolar depression, including emotional processing and social cognition(Gillissie et al., 2022; Takahashi et al., 2010). Interestingly, cortical abnormalities in the temporal lobe have been commonly described in MDD and BD (Hibar et al., 2018; Schmaal et al., 2017). Furthermore, an accumulating body of evidence shows that the neurobiology of depression involves structural and functional dysfunctions in the corticolimbic network, particularly in the temporal lobe, hippocampus, and amygdala (Tassone et al., 2022; Zhong et al., 2016). Recently, Leaver et al. (2022) hypothesized a “network mechanism” of ECT, according to which seizures originating from the middle temporal lobe improve depressive symptoms by correcting or resetting corticolimbic alterations (Leaver et al., 2022). It has been suggested that the increase in temporal CT after ECT could be due to brain damage, including reactive inflammation or edema due to the electrical current near the site of electrode application (Coffey et al., 1991); thus, an increase in temporal CT after ECT could be considered a transient epiphenomenon of treatment, rather than a mechanism underlying its efficacy (Puri et al., 1998). However, although some accumulation of extracellular fluid has been observed using diffusion tensor imaging (Repple et al., 2020), most studies have not detected post-ECT gliosis or brain edema (Szabo et al., 2007), and specific studies, using T2-relaxometry to detect significant fluid shifts, did not support edema as a cause of structural changes detected after ECT (Girish et al., 2001; Kunigiri et al., 2007; Yrondi et al., 2018). Furthermore, a systematic review and meta-analysis of the effects of ECT on brain volumes (Gbyl and Videbech, 2018) did not support the theory of brain damage and did not find an increase in brain damage markers (Agelink et al., 2001; Kranaster et al., 2014; Zachrisson et al., 2000). These findings were confirmed by Gylb et al. (2022), who found no differences in serum S100B, a marker of brain injury, after an ECT series (Gbyl et al., 2022). Crucially, cortical thickening in the temporal lobe in MDD has been previously reported in association with antidepressant drug treatment (Nemati and Abdallah, 2020), suggesting that these effects could be a common mechanism underlying the effectiveness of antidepressant treatments. Consistent with the theories of neurotrophic and neuroplasticity of depression (Duman and Li, 2012), the observed changes in CT during ECT treatment may be the result of underlying increased neural growth, synaptogenesis, and synaptic rearrangement. This is consistent with an increase in neurotrophins after ECT [62]. Nonetheless, further confirmation is needed through randomized studies.

Increased CT in the superior and inferior temporal gyrus was associated with an improvement in depressive symptoms only in one study (Pirnia et al., 2016). Furthermore, Gbyl et al. (2019) observed that although CT in the temporal lobe increased after the first ECT, it returned to baseline after six months, even if the antidepressant effect persisted (Bolwig, 2011).

Taken together, this evidence suggests that temporal CT is primarily affected by ECT, regardless of electrode placement. Less clear is the role of temporal CT changes in clinical improvements. Although most investigations showed that changes in CT did not correlate with clinical symptoms, this was not replicated in all included studies. Therefore, more evidence is needed to clarify the role of temporal CT in changes in depressive symptoms.

### Insula

Seven investigations reported an increase in CT in the insula after ECT (Bracht et al., 2023; Gbyl et al., 2019; Gryglewski et al., 2019; Ji et al., 2022; Pirnia et al., 2016; Sartorius et al., 2016; van Eijndhoven et al., 2016; Xu et al., 2019). Previous literature has shown that the insula contributes to emotion regulation and pain perception and is also involved in the pathophysiology of MDD and bipolar depression (Nagai et al., 2007; Qiu et al., 2020; Schmaal et al., 2017; Sprengelmeyer et al., 2011). A study showed a correlation between cortical thickening in this region and reduced severity of depression (van Eijndhoven et al., 2016). In particular, Bracht et al. (2023) found an increase in CT in the right insula in both ECT and TAU responders, suggesting a potential role for CT as a marker of treatment response, which may not be specific for ECT (Bracht et al., 2023). In addition, recent evidence has shown that insular metabolic activity may represent a potential predictive biomarker of remission in MDD after treatment with antidepressants or psychotherapy (Kelley et al., 2021). Overall, this literature suggests that structural and functional insular neuroimaging abnormalities may play a role as biomarkers of antidepressant effects and predictive biomarkers of symptom remission, regardless of the type of treatment. In summary, the study results suggest that although ECT is associated with an increase in insular CT, the improvement in depression symptoms after ECT does not appear to be associated with CT changes in the insula.

### Frontal lobe

The frontal lobe, in particular the OFC, the superior, middle, and inferior frontal gyrus, have been consistently implicated in the pathophysiology of unipolar and bipolar depression (Altshuler et al., 2008; Drevets, 2007; Fitzgerald et al., 2008). Three of the ten studies included in the present review showed cortical thickening in these areas (Bracht et al., 2023; Gbyl et al., 2019; Xu et al., 2019) and, interestingly, only in the study by Xu et al. (2019) the electrodes were localized in the frontal lobe. Remarkably, Gbyl et al. (2019) reported a strong association between cortical thickening of the right lateral OFC and the antidepressant effect of ECT (Gbyl et al., 2019; Schmitgen et al., 2020). In depression, a reduced volume along with altered activity in OFC has previously been reported (Drevets, 2007). A recent study showed that direct stimulation of this region produced an acute improvement in depressive symptoms in epileptic subjects with moderate to severe depression (Rao et al., 2018). Noticeably, OFC CT increased immediately after the ECT series but returned to baseline levels at six months of follow-up, despite the continued antidepressant effect, suggesting that the persistence of the clinical efficacy of ECT was not related to orbitofrontal CT. Interestingly, a recent GEMRIC consortium study identified a multivariate discriminative pattern of volume alterations in the cortical midline, striatal, and lateral prefrontal areas that differentiated responders from non-responders, suggesting that structural changes in prefrontal areas might play a role in the clinical response to ECT (Mulders et al., 2020).

### Other brain areas

ECT was associated with cortical thickening in several other areas, including the ACC, the parahippocampal gyrus, the entorhinal cortex, the fusiform gyrus, the superior and inferior parietal gyrus, the pre- and postcentral gyrus, and the lateral occipital gyrus. Although all these areas have been consistently implicated in the pathogenesis of unipolar and bipolar depression and appear to be influenced by antidepressant treatments (Bartlett et al., 2018; Klimes-Dougan et al., 2018; Yi et al., 2022), only increased CT in the right lateral occipital gyrus, an area involved in the perception of the face and emotion (Nagy et al., 2012), was associated with clinical improvement (Xu et al., 2019).

### Biological significance of CT findings

Cortical thickening has been previously reported in patients with depression after antidepressant medications (Bartlett et al., 2018; Koenig et al., 2018; Krause-Sorio et al., 2020) and neurostimulation treatments (Dalhuisen et al., 2021; Phillips et al., 2015; Wu et al., 2022), suggesting that cortical changes may not be specifically associated with ECT, but rather represent a generally shared mechanism of effectiveness of various antidepressant treatments. Surprisingly, however, both pretreatment CT and the early increase in CT seem to represent predictive biomarkers of the clinical response to antidepressants(Phillips et al., 2015) suggesting that various processes can underlie its clinical effectiveness. Neuroplasticity, including the processes of neurogenesis, synaptogenesis, dendrogenesis, gliogenesis, and angiogenesis, has been hypothesised to be the biological mechanism for the effects of ECT on CT (Hellsten et al., 2005; Jaggar et al., 2023; Wennström et al., 2004; Zhang et al., 2023). In particular, animal studies have reported that rats subjected to electroconvulsive stimulation show increased genesis of hippocampal neurons with the potential for long-term survival (Altar et al., 2003; Madsen et al., 2005; Olesen et al., 2017). In humans, ECT is associated with a rapid and widespread increase in gray atter volume in the post-treatment phase (Ousdal et al., 2020), as well as an increased brain-derived neurotrophic factor (BDNF) and vascular endothelial growth factor (VEGF) (Sorri et al., 2021; Vanicek et al., 2019). However, the promptness of these volumetric changes suggests that neurogenesis is unlikely to be the only mediating factor for the volumetric effects of ECT (Ousdal et al., 2022). Taking into account this evidence, Ousdal et al. (2022) conceptualized a time-dependent model of the neural effects of ECT that includes three consecutive phases: disruption as the immediate effect, increased neuroplasticity as the short-term effect, and rewiring as the long-term effect (Ousdal et al., 2022). Most of the studies included in our review examined the short-term mechanisms of ECT. The only study that evaluated the long-term effects of ECT reported that CT returned to baseline levels after 6 months (Gbyl et al., 2019), suggesting that CT may not be suitable to detect potential “rewiring” effects. Importantly, the findings of broad cortical thickening after ECT are in line with volumetric studies (Ousdal et al., 2022). Interestingly, two of the included studies explored Gray Matter Volume (and CT (Gryglewski et al., 2019; Sartorius et al., 2016). Although regional structural changes following ECT were detected by both types of measures, they showed different patterns, suggesting that each measure could reflect specific histological changes. We hypothesize that CT can be more sensitive in detecting changes associated with brain atrophy (Lemaitre et al., 2012) compared to gray matter volume morphometry, that can better measure neuroplasticity and rewiring processes after electrical stimulation (Argyelan et al., 2019). Therefore, structural measures may provide complementary information that could help to understand the mechanisms underlying the differential changes in the brain across the cortical regions after ECT.

### Limitations and conclusions

The findings of this review should be interpreted in light of the limitations of primary studies. First, only six studies had a control group, and not all studies reported baseline differences in CT between patients and HC. In addition, most studies did not have a randomized design, thus reducing the strength of evidence supporting a causal link between ECT and changes in CT. Furthermore, the sample sizes were relatively small. Second, concurrent medications (e.g., antidepressants) could also have contributed to CT changes, and this effect could only be estimated in the few controlled studies. Third, the different clinical phenotypes of the participants (e.g., comorbidities, illness phase, duration of the illness) may have contributed to the heterogeneity of the results. In addition, two studies also included individuals with bipolar depression, which may have affected the results. Fourth, in this literature, the sex-related difference in CT changes after ECT was not investigated, and this could be due to the lack of evidence of sex differences in CT in depression. Lastly, the heterogeneity in the positioning of the electrodes during the administration of the ECT may have also influenced CT results.

In conclusion, available studies lend support to the hypothesis that ECT in depression is associated with cortical thickening of the temporal lobe, insula, and, to a lesser extent, frontal lobe, and several other areas of the brain involved in the pathophysiology of depression. These findings are consistent with evidence of increased CT after pharmacological and neuromodulation treatments, suggesting that cortical changes may not be specific to ECT, but rather reflect mechanisms of therapeutic response. Future studies with a randomized design, longer follow-up periods, and multiple methodologies of assessment of morphologic changes are warranted to increase knowledge of the effectiveness of ECT. Such studies may also clarify the potential role of CT and other cortical measures, including surface area, gyrification, and cortical complexity, as biomarkers of the clinical response in depression.

## Data Availability

All data produced in the present work are contained in the manuscript

## Acknowledgments

None.

