## Supplementary material for "Electroconvulsive therapy increases cortical thickness in depression: A systematic review"

*Assessment of methodological quality*

The methodologcial quality of the included studies was assessed using a point system derived from Gbly and Videbech (2018) (Gbyl and Videbech, 2018). The quality score consists of the following items: a) number of subjects (0,1 point was assigned for every subject enrolled); b) the presence of a control group (1 point was assigned for the presence of an age- and gender-matched group of healthy controls, additional 1 point for the presence of a control group of depressed patients not treated with ECT); c) the number of MRI scans the control group underwent (1 point assigned if the group was scanned twice or more); d) MRI scanner field strength (1 point for 3 Tesla); e) voxel size (1 point given for voxel size lower than 1.0 mm^3^; f) medication status (1 point if subjects were not medicated or if medication had been washed out in >80% of subjects before inclusion); g) consecutively collected sample (1 point was assigned if it was explicitly stated that a sample was collected in a consecutive way); h) duration of follow-up time (1 point for every MRI scan conducted later than 2 weeks after completion of the ECT series). The higher the score, the better the methodological quality.

**Table S.1. Assessment of the methodological quality of the included studies.**

| **Study** | **N° of subjects** | **Control group** | **N° of MRI scans in the control group** | **MRI scanner field strength** | **Voxel size** | **Medication status** | **Consecutively collected sample** | **Duration of follow-up** | **Total quality score** |
| --- | --- | --- | --- | --- | --- | --- | --- | --- | --- |
| Pirnia et al., 2016 | 2.9 | 1 | 1 | 1 | 0 | 1 | 0 | 0 | 6.9 |
| Sartorius et al., 2016 | 1.8 | 1 | 0 | 1 | 0 | 0 | 0 | 0 | 3.8 |
| Van Eijndhoven et al., 2016 | 2.3 | 1 | 1 | 0 | 0 | 1 | 0 | 0 | 5.3 |
| Gbyl et al., 2019 | 1.8 | 0 | 0 | 1 | 1 | 0 | 0 | 1 | 4.8 |
| Gryglewski et al., 2019 | 1.4 | 0 | 0 | 1 | 0 | 0 | 0 | 0 | 2.4 |
| Schmitgen et al., 2019 | 1.2 | 1 | 0 | 1 | 0 | 0 | 0 | 0 | 3.2 |
| Xu et al., 2019 | 2.3 | 0 | 0 | 1 | 0 | 0 | 0 | 0 | 3.3 |
| Yrondi et al., 2019 | 1.7 | 1 | 1 | 1 | 0 | 0 | 0 | 0 | 4.7 |
| Bracht et al., 2023 | 2.0 | 2 | 1 | 1 | 0 | 0 | 0 | 0 | 6 |
| Ji et al., 2023 | 5.6 | 1 | 1 | 1 | 0 | 0 | 0 | 0 | 8.6 |
